# ANALYSIS OF MYELOSUPPRESSION IN POST-CHEMOTHERAPY BREAST CANCER PATIENTS (A Study at Division of Hematology and Medical Oncology Airlangga University Hospital)

**DOI:** 10.1101/2024.01.31.24302087

**Authors:** Yulistiani, Febriansyah Nur Utomo, Pradana Zaky Romadhon, Andwynanda Bhadra Nareswari

## Abstract

**Backgrounds:** Myelosuppression is decreased bone marrow activity that can be caused by chemotherapy which lowers the formation of blood cells and manifests as anaemia, neutropenia, leukopenia, and thrombocytopenia.

**Objective:** This study aims to analyze the post-chemotherapy myelosuppression in breast cancer patients and the chemotherapy regimen used during chemotherapy associated with blood parameters.

**Method:** This study was conducted in observational retrospective design. Data were taken from patients’ medical record that went on chemotherapy during January 2021-December 2022.

**Results:** The result showed that 97% patients had myelosuppression with the significance statistic’s analysis P<0,05. The chemotherapy regimens used in breast cancer patients are AC-T, TAC, Paclitaxel-Carboplatin, TC, AC-TH, and TCH. This study found myelosuppression in several regimens: anaemia grade I-II in patients with AC-T regimen, neutropenia grade I-IV in patients with TCH regimen, leukopenia grade I-IV in patients with TC regimen, and thrombocytopenia grade I in patients with Paclitaxel-Carboplatin regimen. The incidence of delayed chemotherapy in patients due to myelosuppression is 18%.

**Conclusion:** Based on this study, all the regimens used had myelosuppression side effect. The highest incidence of myelosuppression occurred in patient with TAC and TCH regimen. Myelosuppression can cause delayed therapy, dose reduction, and drug substitution.

## Introductions

The definition of myelosuppression according to the National Cancer Institute in 2022 is a condition in which bone marrow activity decreases which causes the formation of red blood cells, white blood cells, and platelets to decrease. Myelosuppression is a chemotherapy complication that often occurs due to damage to hematopoietic stem cells and progenitor cells in the bone marrow, and most often manifests as anaemia, neutropenia, and thrombocytopenia (Epstein, 2020). Chemotherapy can suppress the hematopoietic system and damage the host cell protection mechanisms and cause myelosuppression (Carey, 2003). Grading criteria for hematologic toxicity caused by chemotherapy can be seen on **Table 1**

**Table 1.** Grading Criteria for Hematologic Toxicity Adverse Effect (National Cancer Institute, 2017)

| Adverse Effect | Unit of Measure | Grade 1 | Grade 2 | Grade 3 | Grade 4 | Grade 5 |
| --- | --- | --- | --- | --- | --- | --- |
| Anaemia | Haemoglobin (g/dl) | Less than LLN - 10 | <10 - 8 | < 8 | Life threatening; urgent intervention indicated | Death |
| Neutropenia | Absolute Neutrophil Count ($\times 10^9$) | Less than LLN - 1,5 | 1 - 1,5 | 0,5-1 | <0,5 | Death |
| Thrombocytopenia | Platelet count ($\times 10^9$) | Less than - 75 | 50 - 75 | 25 - 50 | <25 | Death |
| Leukopenia | Leukocyte count (/mm<sup>3</sup>) | Less than LLN - 3000 | <3000 – 2000 | <2000 – 1000 | <1000 | Death |

Clinical meta-analysis studies show that the incidence of myelosuppression in breast cancer patients receiving anthracycline-based chemotherapy in western countries is around 80% (Cui L, *et al*., 2021). In Asian early-stage breast cancer patients receiving anthracycline regimens, the incidence of grade 4 neutropenia and febrile neutropenia was 44.6% and 8.5% (Kim HS, *et al*., 2016). In breast cancer patients who experience myelosuppression while undergoing chemotherapy the incidence of neutropenia can reach 16.7%, the incidence of anaemia reaches 17% with Adriamycin and cyclophosphamide chemotherapy regimens, and the incidence of thrombocytopenia reaches 20% -25% in the first cycle of chemotherapy (Tia L, *et al*, 2015).

Drug-induced myelosuppression is a side effect of myelosuppression that depends on the type of drug and dose for cancer therapy. Bone marrow contains stem cells that are capable of reproducing and differentiating into red blood cells, white blood cells, and platelets. Cells produced by stem cells are almost always in mitosis and reproduce rapidly, making them very susceptible to cytotoxic damage. Cytotoxic chemotherapy can suppress the hematopoietic system and damage the protective mechanisms of host cells (Carey, 2003). Anthracycline-based regimen therapy is still the standard first-line regimen for breast cancer (Jasra S and Anampa J., 2018). Cytotoxic drugs that can be used alone or in combination as chemotherapy for breast cancer include doxorubicin, cyclophosphamide, paclitaxel, docetaxel, and vincristine (Fernando A, *et al*., 2021).

Doxorubicin can induce myelosuppression through the metabolic processes of reactive oxygen species or ROS directly by inducing platelet cytotoxicity by stimulating the formation of ROS. Potentiation of ROS production and reduced endogenous antioxidants can trigger the intrinsic apoptotic pathway in hematopoietic cells (Chen T, *et al*., 2017). Cyclophosphamide can induce oxidative stress in the bone marrow which results in suppression of anti-oxidative enzymes and causes myelosuppression. Paclitaxel causes myelosuppression by binding to microtubules of bone marrow cells, thereby preventing depolymerization. This process then causes inhibition of mitosis and then apoptosis of cell division. New findings on ROS have shown that drugs with a ROS mechanism are used to treat cancer because they accelerate tumor cell death. High levels of ROS induce not only tumor cell death but also oxidative damage to normal cells, especially haemopoietic cells in the bone marrow, leading to bone marrow suppression and other adverse effects (Fernando, 2021).

The incidence of myelosuppression can be observed by clinical examination and laboratory examination of a complete blood count (Madeddu C, *et al*., 2018). The decrease in blood cell count did not occur immediately after cytotoxic therapy. The lowest blood cell count after chemotherapy treatment is called the nadir, and each type of blood cell has a different nadir. The number of red blood cells will decrease on the 7-10th day after chemotherapy. White blood cells and platelets reach nadir 7-14 days after chemotherapy. The time needed for recovery of platelets, neutrophils, and hemoglobin is 2-7 days, 7-8 days, and 7-14 days during the first cycle of chemotherapy (Norman H, *et al*., 2022). In patients who experience degree 4 neutropenia, the neutrophil value will decrease on the 10-15th day after chemotherapy (Tia L, *et al*, 2015).

Some patients recover rapidly from acute myelosuppression after chemotherapy, but there are cases of residual myelosuppression that persists for a long time and manifests as decreased reserves and impaired renewal of the hematopoietic stem cells. The occurrence of long-term myelosuppression was more common in patients receiving treatment with carboplatin, busulfan, bischloronitrosourea and/or total body irradiation, and very rarely in patients receiving single injections of relatively less cytotoxic agents (Wang Y, *et al*., 2006).

Myelosuppression causes delays, reductions, and discontinuation of doses which will affect the reduction of the intensity of chemotherapy doses and can jeopardize the effectiveness of therapy (Epstein, 2020). Dosage reduction may be considered empirically before chemotherapy if the patient has a low initial neutrophil or platelet count, has reduced bone marrow reserves, has impaired drug elimination, or is receiving a combination of agents that induce myelosuppression (Epstein, 2021). To prevent the side effects of myelosuppression, primary prophylaxis can also be used with Granulocyte-Colony Stimulating Factor (G-CS) F in neutropenic patients or an erythropoiesis-stimulating factor, namely epoetin in anemic patients (Carey, 2003). Some chemotherapy regimens use myeloid growth factors (MGFs) to help increase blood cell counts and prevent infection (Gradishar WJ, *et al*., 2022).

Routine complete blood count needs to be done, both before and after chemotherapy cycles, as an initial examination, to identify signs or symptoms of myelosuppression side effects. Myelosuppression is a side effect that is a major concern in therapy. In cases of grade 3-4 (severe) myelosuppression, the patient does not receive chemotherapy beforehand (delayed) until the myelosuppression improves. In this case, the prevention and treatment of myelosuppression is very important. Treatment for preventing myelosuppression can be in the form of G-CSF prophylaxis and for treatment of myelosuppression therapy can be in the form of blood transfusions if needed, administration of epoetin in anaemia.

If anaemia occurs, the cells in the body do not get enough oxygen so they tire easily, the skin becomes pale, and headaches (Gradishar WJ, *et al*., 2022). A reduced number of neutrophils can cause immunosuppression so that the body’s ability to fight infection will decrease (Gradishar WJ, *et al*., 2022).

With this study, it can be known the type of chemotherapy that causes myelosuppression, the effects of myelosuppression, the number of delays in chemotherapy cycles, and the management of this side effect. This study can be used as a means of monitoring and evaluating the quality of pharmaceutical services.

## Method

### Patients and data collection

This observational study was conducted in a retrospective design to observe and analyzed myelosuppression side effect post chemotherapy in breast cancer patients using blood parameters and associated with the regimen used. Data were taken from medical records of all patients who were diagnosed with breast cancer and went on chemotherapy with complete blood counts test including hemoglobin, neutrophil, platelet, and leukocyte counts during January 2021 - December 2022. The study instrument used in this study was a data collection sheet from patients’ chemotherapy medical records. The inclusion criteria in this study were patients diagnosed with all breast cancer and undergoing chemotherapy with complete blood counts (hemoglobin, neutrophil, platelet, and leukocyte counts) and data on drug regimens and doses used during chemotherapy and data on post-chemotherapy side effects. There were no exclusion criteria for this study. All patients’ data for this study were collected from March 10^th^ – June 30^th^ 2023 and remain anonymized before and after data collection. The study was approved by the Research Ethics Committee of Rumah Sakit Universitas Airlangga and was declared to have passed the ethical review with a Certificate of the Ethics Review Approval Number 021/KEP/2023.

### Data analysis

Presentation of patient demographic data (gender, age, and cancer stage), chemotherapy regimen used, as well as myelosuppression incidence each cycle in certain chemotherapy regimen were presented descriptively in the form of frequency (n) and percentage (%). Data analysis using statistics was used to determine the difference of blood parameters before and after chemotherapy. The data obtained were tested for normality. If the results of the normality test showed that the data was normally distributed (sig. >0.05), then the data would be tested using the T-paired test. If the results of the normality test showed that the data was not normally distributed (sig. <0.05), then the data would be tested using the Wilcoxon method.

### Results

In total, the data were taken from 35 patients; all breast cancer patients undergoing chemotherapy were all female. This is in accordance with statistical data conducted by the American Cancer Society in 2022, reporting that more women experience breast cancer than men. However, breast cancer can also be experienced by men. The age distribution of patients was divided into two groups based on the average time of menopause, namely age under 50 years and age above 50 years (CDC, 2022). The risk of breast cancer increases with increasing age of patients over 50 years. From the results, 66% of patients diagnosed with breast cancer were over 50 years old, the youngest patient was 32 years old and the oldest patient was 72 years old. These results are the same as the epidemiological study above. In the following discussion, tables and figures, the data will be processed based on the number of patients who suffer from the side effect of myelosuppression which was 34 (97%) patients. Patients’ characteristics are summarized on **Table 2**

**Table 2.**
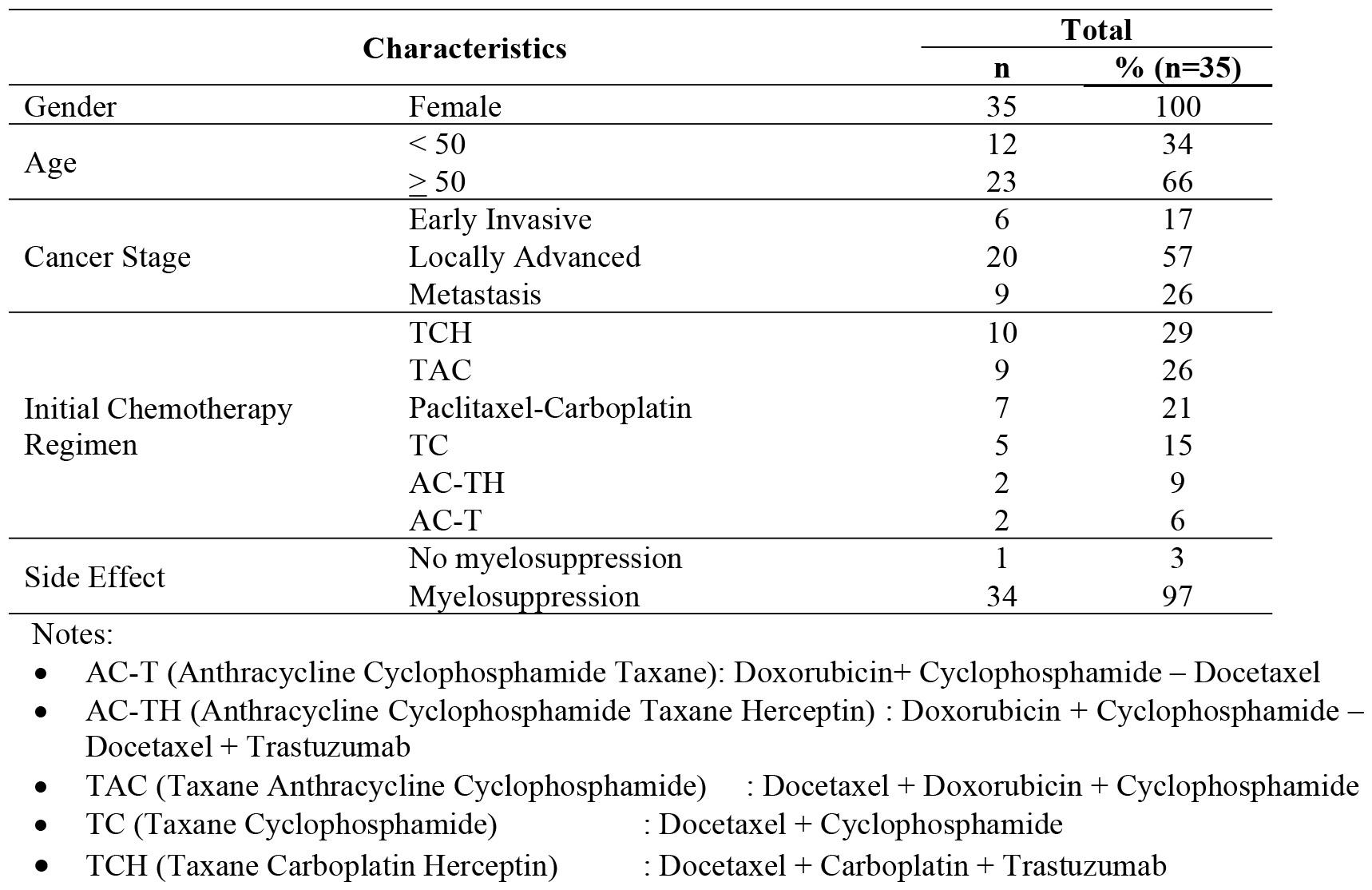
Patients Characteristics.

Patients might have regimen changes during chemotherapy from the planned initial regimen (**Table 3**). Changes in chemotherapy regimens could occur due to the occurrence of certain side effects or the regimen previously used did not cause an adequate response to the cancer cell. Chemotherapy regimen changes might occur one or more times during chemotherapy. Changes in chemotherapy regimens experienced by patients in this study were in accordance with guidelines published from NCCN, 2018 which described several regimens that could be recommendations or alternatives in administering chemotherapy according to the patient’s condition.

**Table 3.** Changes in Regimen during Chemotherapy.

| Initial Chemotherapy Regimen | Regimen Chemotherapy After Changes | Chemotherapy Cycle | n |
| --- | --- | --- | --- |
| Paclitaxel-Carboplatin | TAC | 4 | 1 |
| TAC | AC-TH | 5 | 1 |
|  | TCH | 5 | 3 |
| TC | AC-T | 3 | 1 |
| **Total** |  |  | **6** |

Notes :
- n : number of patients

**Table 3.**
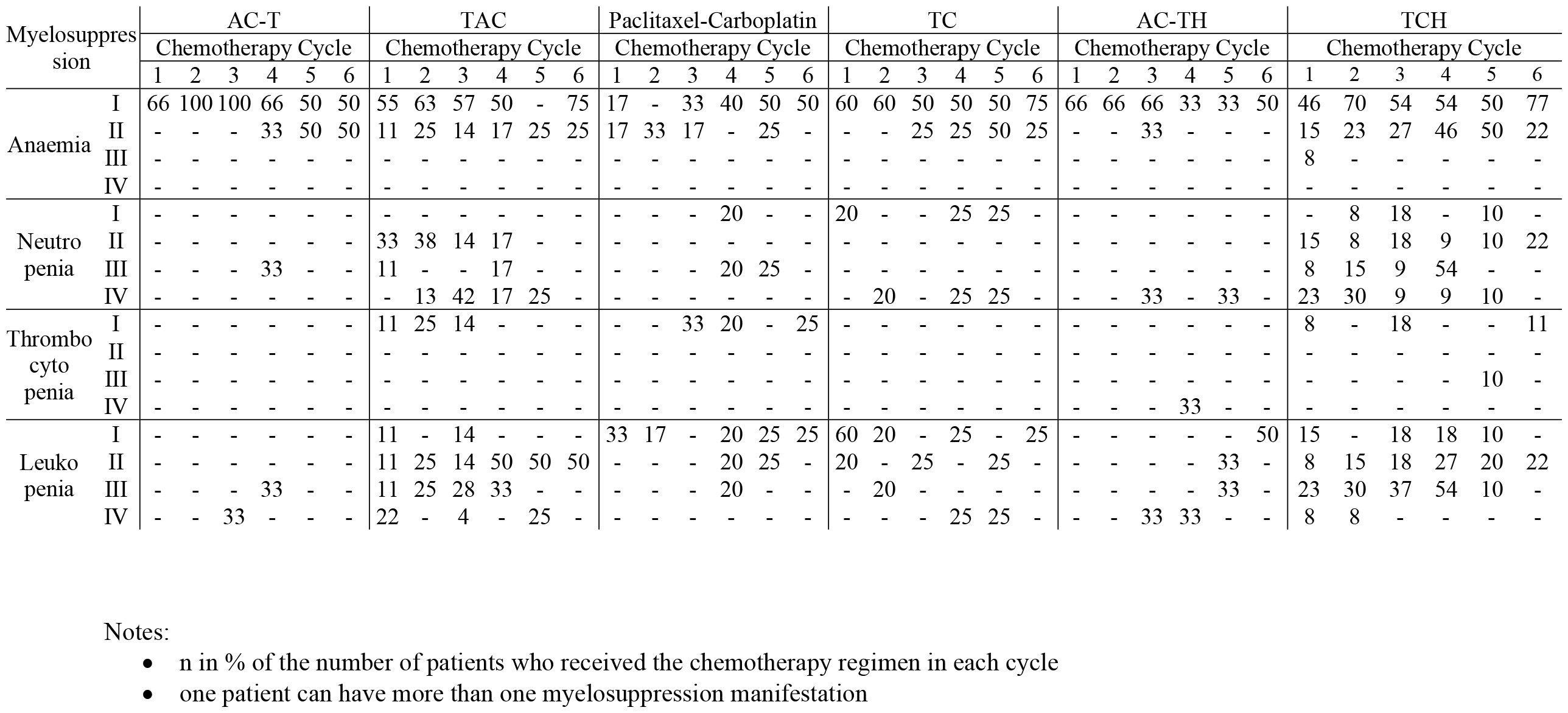
Percentage of Patient with Myelosuppression from 1^st^ to 6^th^ Cycle Chemotherapy and Chemotherapy Regimen.

Breast cancer patients who had chemotherapy will undergo a blood sampling procedure to see the number of patient’s blood conditions both before and after chemotherapy. Results of blood count data obtained after chemotherapy then grouped into degrees of myelosuppression based on CTCAE 2013 and 2017. **Figure 1** shows that the most common side effect of myelosuppression experienced by patients was anaemia in each cycle. The number of anemic patients increases along with the continuation of the cycle. The increase in the number of patients with anaemia could be caused by the cumulative risk of anaemia, so that the number of anaemia patients increase with increasing cycles of chemotherapy. Then, it was followed by the number of patients who experienced side effects leukopenia and neutropenia. The number of patients who experienced leukopenia and neutropenia were increased and decreased during chemotherapy this could be associated with the therapy given to patients. The patient was given filgrastim injection after chemotherapy on day three as a form of prevention and treatment to increase white blood cells and neutrophils. With the administration of filgrastim injection on the 3rd day after chemotherapy and a complete blood count after chemotherapy was carried out on the 6th day, there were several variations because of the filgrastim therapeutic outcome. Thrombocytopenia side effects occurred in smaller numbers than the other three side effects. Patients who experienced thrombocytopenia were more in grade I and one patient had grade III thrombocytopenia. In this study, patients might have more than one manifestation of myelosuppression in one cycle.

**Figure 1.**
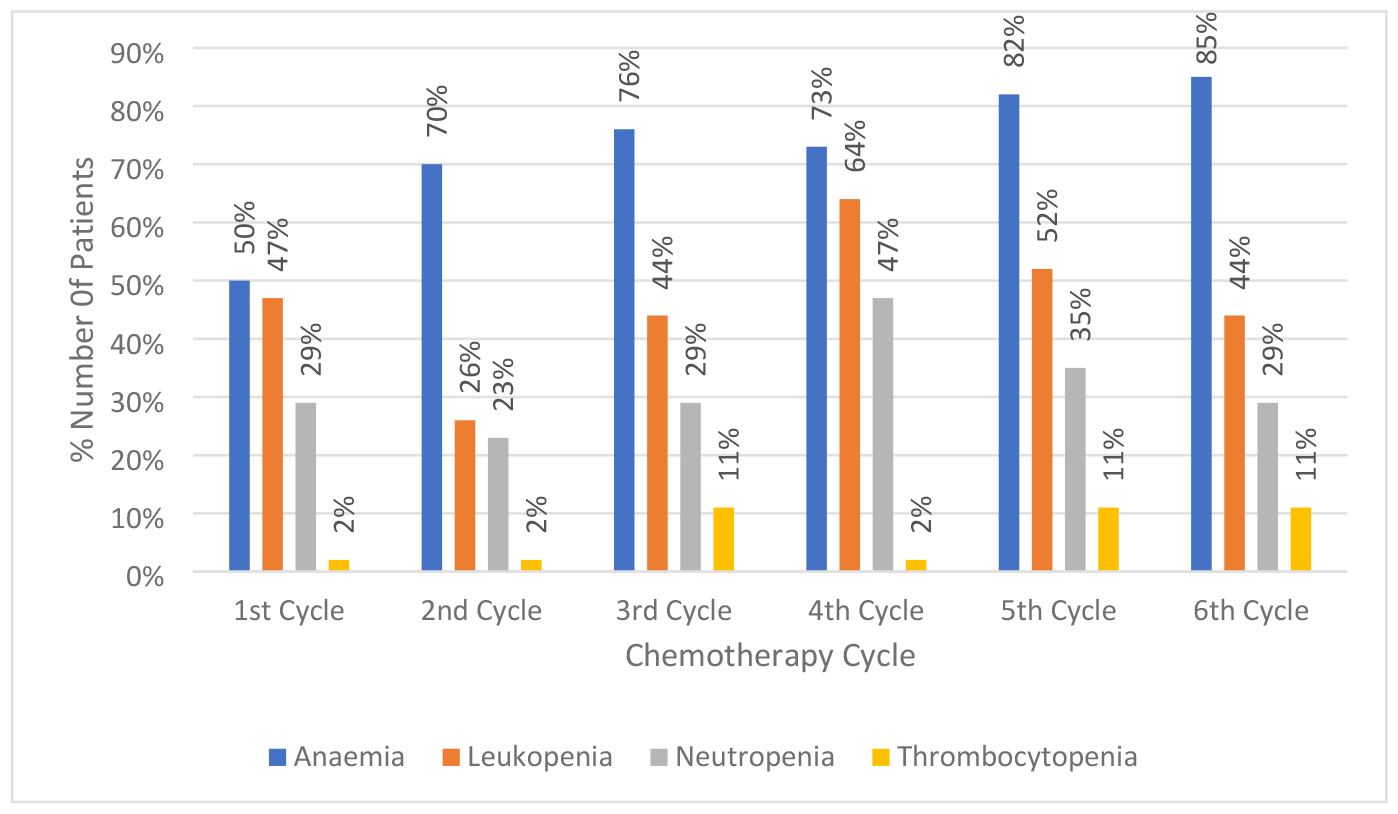
Patient’s Chemotherapy Cycle Profile.

Patients who received the same chemotherapy regimen were grouped into one regimen group to see the number of the myelosuppression side effect that occurred in each regimen. In each cycle, various side effects can occur with varying severity. Patients who experienced a change in chemotherapy regimen and experience myelosuppression would be grouped together with the new regimen group and in cycle 1 again. Table 3 contains data showing the number of occurrences of myelosuppression side effects and their severity grading. The occurrence of myelosuppression was also associated with the chemotherapy regimen used.

Based on the data obtained and then tabulated according to **Table 3**, it resulted that in the AC-T regimen there were two patients who underwent complete chemotherapy cycles according to the initial regimen and one patient who underwent a regimen change to AC-T. In the AC-T regimen, the manifestations of myelosuppression found were anaemia in degrees I-II, neutropenia in degrees III, and leukopenia in degrees III-IV. No thrombocytopenia was found in patients receiving the AC-T regimen. In the TAC regimen, there were four patients who underwent six complete chemotherapy cycles according to the initial regimen, one patient underwent a regimen change to TAC, and four patients received the initial TAC regimen and then changed to another regimen. In the TAC regimen, the manifestations of myelosuppression found were anaemia in degrees I-II, neutropenia in degrees II-IV, thrombocytopenia in degrees I, and leukopenia in degrees I-IV. In the Paclitaxel-Carboplatin regimen, there were four patients who underwent six complete chemotherapy cycles according to the initial regimen and two patients received the initial Paclitaxel-Carboplatin regimen and then changed to another regimen. In the Paclitaxel-Carboplatin regimen, there were several manifestations of myelosuppression found, anaemia of I-II degrees, neutropenia of I and III degrees, thrombocytopenia of I degrees, and leukopenia of I-III degrees. In the TC regimen there were two patients who underwent 6 complete chemotherapy cycles according to the initial regimen and one patient received the initial TC regimen and then changed to another regimen. In the TC regimen, the manifestations of myelosuppression found were anaemia in degrees I-II, neutropenia in degrees I and IV, and leukopenia in degrees I-IV. No thrombocytopenia was found in patients receiving the TC regimen. In the AC-TH regimen, there were two patients who underwent six complete chemotherapy cycles according to the initial regimen and 1 patient who underwent a regimen change to AC-TH. In the AC-TH regimen, the manifestations of myelosuppression found were grade I-II anaemia, grade IV neutropenia, grade IV thrombocytopenia, and grade I-IV leukopenia. In the TCH regimen, there were nine patients who underwent six complete chemotherapy cycles according to the initial regimen, three patients underwent a regimen change to TCH, and one patient received the initial TCH regimen and then changed to another regimen. In the TCH regimen, the manifestations of myelosuppression found were grades I-III myelosuppression, neutropenia degrees I-IV, degrees I and III thrombocytopenia, and degrees I-IV leukopenia.

A statistical test (t-test) was ran to test whether myelosuppression effect that caused by chemotherapy is statistically significant compared to the baseline data before chemotherapy (**Table 4**), Based on the results of the different test, it was found that the data had significant differences and insignificant differences. If the P value> 0.05, there is no significant difference between the blood data before and after chemotherapy. Conversely, if the P value <0.05, there is a significant difference between the blood data before and after chemotherapy. If there is a significant difference between the data before and after chemotherapy, it can be concluded that the chemotherapy drugs used reduce blood parameters and the decrease is significant. All blood parameters in the TC and TCH regimens experienced significant differences which showed that the drugs in this regimen caused a decrease in blood parameters (**Table 4**). In the TAC and Paclitaxel-Carboplatin regimens, the ANC, platelets, and leukocyte parameters showed significant differences. In the AC-TH regimen, only the Hb and platelet parameters had significant differences. Whereas in the AC-T regimen, it was only the Hb parameter that has a difference.

**Table 4.** Statistics Test on Pre-Post Chemotherapy Blood Parameters.

| Chemotherapy Regimen | Blood Parameter | Pre-Chemo Mean ± SD | Post-Chemo Mean ± SD | P (significance) |
| --- | --- | --- | --- | --- |
| AC-T | Hb | 11,4 ± 1,2 | 10,7 ± 1,0 | 0,012<sup>a</sup> |
| | ANC | 4714 ± 2046 | 5543 ± 2578 | 0,067<sup>b</sup> |
| | Thrombocyte | 401625 ± 106138 | 372937 ± 726164 | 0,273<sup>b</sup> |
| | Leucocyte | 7235 ± 2077 | 7174 ± 2710 | 0,895<sup>b</sup> |
| TAC | Hb | 11,5 ± 1,5 | 11,4 ± 1,5 | 0,725<sup>b</sup> |
| | ANC | 4366 ± 2155 | 2848 ± 2479 | 0,000<sup>a</sup> |
| | Thrombocyte | 301750 ± 73935 | 206000 ± 50661 | 0,000<sup>a</sup> |
| | Leucocyte | 5865,5 ± 2418 | 4338 ± 5064 | 0,000<sup>a</sup> |
| Paclitaxel Carboplatin | Hb | 11,7 ± 1,3 | 11,9 ± 1,7 | 0,369<sup>b</sup> |
| | ANC | 5952 ± 3155 | 3889 ± 1490 | 0,043<sup>a</sup> |
| | Thrombocyte | 309411 ± 92345 | 248235 ± 81181 | 0,003<sup>a</sup> |
| | Leucocyte | 6683 ± 3248 | 5045 ± 2014 | 0,014<sup>a</sup> |
| TC | Hb | 11 ± 2,3 | 11,6 ± 2,2 | 0,029<sup>a</sup> |
| | ANC | 5371 ± 2916 | 2835 ± 1500 | 0,001<sup>a</sup> |
| | Thrombocyte | 414185 ± 139901 | 298888 ± 67375 | 0,000<sup>a</sup> |
| | Leucocyte | 8038 ± 3193 | 5044 ± 2429 | 0,000<sup>a</sup> |
| ACTH | Hb | 11,4 ± 2,3 | 11,6 ± 2,2 | 0,018<sup>a</sup> |
| | ANC | 4794 ± 3060 | 3886 ± 2366 | 0,343<sup>b</sup> |
| | Thrombocyte | 480874 ± 81407 | 360000 ± 109688 | 0,000<sup>a</sup> |
| | Leucocyte | 7067 ± 3471 | 5320 ± 2701 | 0,097<sup>b</sup> |
| TCH | Hb | 10,7 ± 1,1 | 10,4 ± 1,3 | 0,001<sup>a</sup> |
| | ANC | 6668 ± 2464 | 1622 ± 1317 | 0,000<sup>a</sup> |
| | Thrombocyte | 321787 ± 130407 | 271106 ± 96426 | 0,000<sup>a</sup> |
| | Leucocyte | 9607 ± 3160 | 3014 ± 1773 | 0,000<sup>a</sup> |

Delays in chemotherapy cycles could be experienced by some patients who experience side effects of myelosuppression. This is known based on the results of laboratory data obtained before starting chemotherapy was too low. Laboratory data results for blood checks that were too low could endanger the patient’s condition because chemotherapy drugs that were cytotoxic not only kill cancer cells, but can also kill cells that divide quickly such as components of blood cells. The occurrence of delays in chemotherapy cycles was undesirable because it could have an impact on the success of therapy. The event of delay was expected to make the patient’s condition ready to get chemotherapy.

It is shown in **Figure 2** that 4 out of 34 (12%) patients experienced one chemotherapy delay and two out of thirty-four (6%) patients experienced two delays in different cycles due to the side effects of myelosuppression. The length of delay of chemotherapy in patients varies from three days to nine weeks.

**Figure 2.**
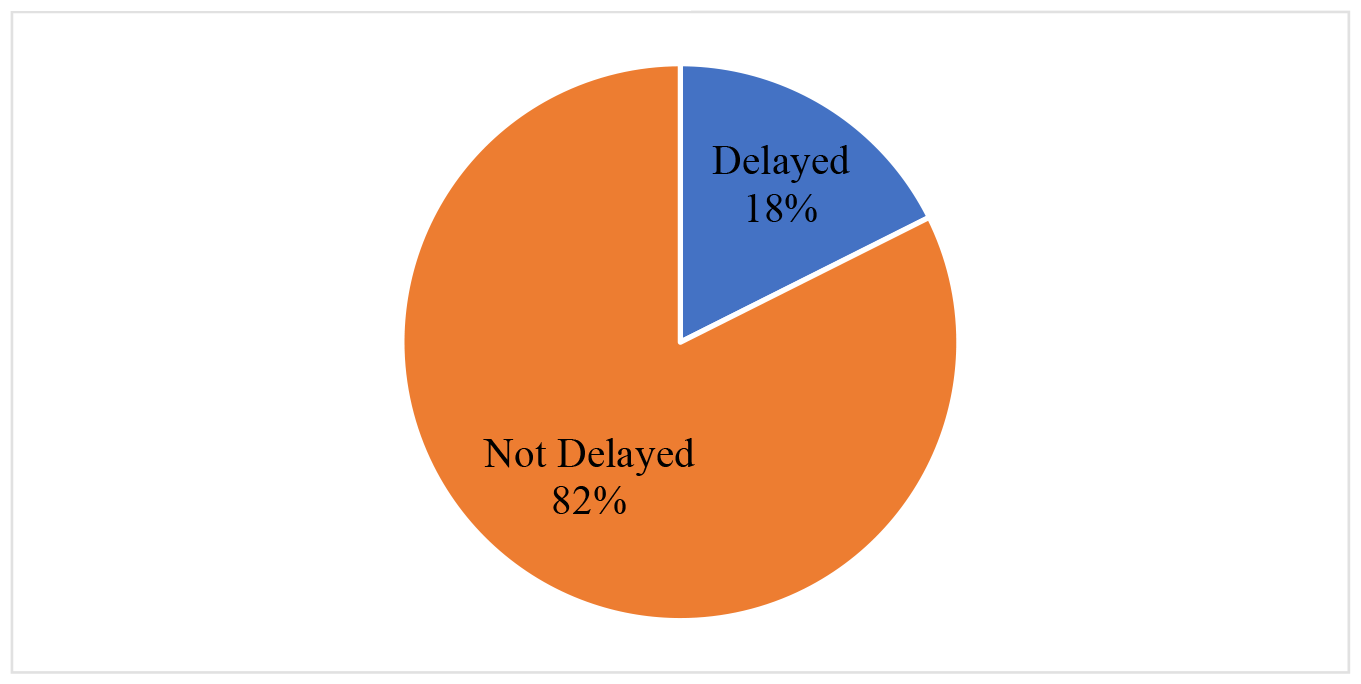
Delay Chemotherapy Profile.

## Discussion

Based on this study, the population of patients diagnosed with breast cancer in 2021-2022 at Airlangga University Hospital was 51 patients. However, of the 51 patients whose medical records were taken, 16 patients did not receive chemotherapy so they did not have complete blood laboratory data and one patient did not experience the side effect of myelosuppression. Patients who did not undergo chemotherapy undergo another therapy modality, namely hormone therapy. From this population, there were 34 patients who met the inclusion criteria. Patients who did not meet the inclusion criteria for chemotherapy clinics are patients who receive hormonal therapy and did not have complete blood count data. Patients undergoing chemotherapy at Airlangga University Hospital did not receive radiation therapy unless referred to undergo radiation therapy at Dr. Soetomo. Based on this study, it can be assumed that all patients affected by the side effects of myelosuppression were the result of the effects of chemotherapy cytostatic drugs.

Patients received chemotherapy regimens according to the stage of the cancer and other conditions such as hormone receptor status. The chemotherapy regimen given to patients at Airlangga University Hospital is the same as in the NCCN guidelines, 2018. In this study, blood parameter data collection was limited to six cycles because most chemotherapy consists of six cycles. However, at the time of data collection, there were several patients who experienced changes in chemotherapy regimens. Changing the chemotherapy regimen to a new regimen will be added to the 1^st^ cycle of the new regimen. If a patient experiences a regimen change from a TAC regimen to an AC regimen, the blood data for the TAC and AC regimens are differentiated and the blood data for the AC regimen will be included in cycle 1 of the AC regimen. So that in **Table 3** the number of patients experienced an increase and decrease in each cycle.

Based on the data obtained, the most frequent manifestation of myelosuppression side effects is anaemia with the number of incidents per cycle being more than 50%, the number of anaemia incidents continues to increase every cycle. This is in accordance with other study which states that anaemia often occurs in cancer patients and is a frequent complication of myelosuppressive chemotherapy (Jerome *et al*., 1999). Of the hematologic disorders, anaemia is found in breast cancer patients (Hu, 2005). The study results showing the increasing incidence of anaemia in patients are also supported by other study which states that the risk of anaemia is cumulative, increasing with increasing cycles of cancer drug treatment. The effects obtained in patients who have anaemia in the first cycle will continue into subsequent cycles and this causes the percentage of patients who experience anaemia to continue to increase. Anaemia in breast cancer patients can occur due to tumour bleeding, tumour invasion of the bone marrow, malnutrition caused by the tumour, abnormal iron metabolism, impaired kidney physiology, and impaired bone marrow function (Candelaria, 2015). The finding of high anaemia side effects in this study is in line with a multicentre study of more than 2800 patients with solid tumours, where the incidence of anaemia increased from 17% before the first chemotherapy cycle to 35% in the sixth cycle (Madeddu C, 2021). The treatment that has been carried out to treat anaemia is giving a PRC transfusion. PRC transfusions are given to patients before chemotherapy. Providing PRC transfusions before chemotherapy is prioritized because the condition for patients undergoing chemotherapy must not be anaemic. If the patient is still anaemic then chemotherapy must be postponed until the patient is not anaemic. The anaemia experienced by post-chemotherapy patients is on average grade 1-II.

Based on data, manifestations of leukopenia have the second largest number of occurrences after anaemia. The number of leukopenia incidents varies in each cycle, decreasing in the 2nd cycle, then increasing in the 3rd and 4th cycles, and decreasing again in the 5th and 6th cycles. The decrease in white blood cell levels begins a few days after chemotherapy is given, reaching the lowest levels in the second or third week after chemotherapy. As the bone marrow recovers from the effects of chemotherapy, the WBC count begins to rise again. The results obtained in this study are like to other studies which reported that the number of white blood cells after chemotherapy decreased significantly in the 4th and 8th cycles and compared with the number of white blood cells before chemotherapy (Aynalem, 2022). The incidence of leukopenia and neutropenia is similar in decreasing and increasing levels of white blood cells and neutrophils. This is because neutrophils are a component of white blood cells, so if the number of white blood cells decreases, the number of neutrophils will also decrease. However, this does not always happen with the same ratio because ANC measurements use the percent of neutrophils, if the percent of neutrophils is high then the number of white blood cells will also be high. The incidence of neutropenia is smaller compared to the incidence of leukopenia. Neutrophils are a type of white blood cell that is an important part of the immune system and helps the body fight infections. To overcome the reduction in white blood cells, filgrastim injection is given to increase the number of white blood cells as immune cells in the body. A decrease in white blood cell parameters can be associated with the use of breast cancer therapy drugs (Nascimento, 2017). Simultaneously, drugs cause damage to normal tissues, including parts of the immune system and immune cells such as lymphocytes, resulting in decreased immunological function (Salako, 2020).

Based on the data obtained, manifestations of thrombocytopenia occur in less than 10% of patients in each cycle. This is in accordance with the results of study conducted by Aynalem (2022) which stated that the number of platelets increased significantly before and after treatment, with a significant increase in the 4th cycle and 8th cycle. This could be due to the ability of cancer cells to cause thrombocytosis and platelet aggregation. Low platelet counts in breast cancer are mostly related to cancer cells which often cause activation of the coagulation system so that platelets are used in large quantities by cancer cells so that circulating platelets decrease in number and manifest as thrombocytopenia. Thrombocytopenia due to chemotherapy occurs due to the type and dose of chemotherapy. Regimens containing gemcitabine, platinum, or temozolomide most commonly cause thrombocytopenia. Bleeding and the need for platelet transfusion in thrombocytopenia are rare except in patients with platelet counts below 25x109/L where the bleeding rate is significantly increased and platelet transfusion is the only treatment.

In the AC-T regimen, the manifestations of myelosuppression found were anaemia in grades I-II as much as 33% -100% in cycles 1 to 6, neutropenia in grade III as much as 33% in cycle 4, and leukopenia in grades III-IV as much as 33% in the 3rd and 4th cycles. No thrombocytopenia was found in patients receiving the AC-T regimen. Whereas in the AC-TH regimen the manifestations of myelosuppression found were anaemia of grade I-II as much as 33% -66% in the 1st to 6th cycles, 33% grade IV neutropenia in the 3rd and 5th cycles, 3rd and 5th degree thrombocytopenia. Grade IV is as much as 33% in the 4th cycle, and leukopenia degrees I-IV as much as 33% - 50% in the 3rd to 6th cycle. This is in accordance with another study conducted by Palukuri NR, (2020) that patients treated with chemotherapy regimen AC reported that 17% of patients had anaemia. In another study, it was also found that the AC regimen could cause FN with a percentage of events ranging from 0.3% to 6% and 6.1% to 25.2%. The AC regimen can be continued with the drug taxane, in the protocol is paclitaxel, for 2 cycles (Palukuri NR, 2020). In the AC-T and AC-TH regimens, the most common side effect experienced is anaemia which can be experienced in cycles 1 to 6 of chemotherapy, so there is a need for vigilance in monitoring anaemia in patients using the AC-T regimen and AC-TH.

The paclitaxel and carboplatin regimen were found to cause myelosuppression side effects with manifestations of anaemia, neutropenia, thrombocytopenia, and leukopenia. As many as 17% - 50% of patients experience anaemia in each cycle with degrees of severity I-II. In other studies, it was found that paclitaxel caused grade 1 or 2 anaemia in 36%-51% of metastatic breast cancer patients (Jerome, 1999). Neutropenia grades I and III occur in 20% - 25% of patients in the 4th and 5th cycles. Leukopenia occurs in 17% - 33% of patients in each cycle except in the 3rd cycle with grades I-III. The paclitaxel-carboplatin regimen is known to have a lower incidence of neutropenia compared to the alternative regimen, docetaxel and carboplatin (Jerome, 1999).

Anaemia caused by the TAC regimen occurred in all cycles with grade I and II severity. It was shown that as many as 11% - 75% of patients experience grade I-II anaemia in all chemotherapy cycles. This is in accordance with the literature which states that the combination of docetaxel, doxorubicin, and cyclophosphamide or the TAC regimen can cause grade 2 and degree 3 anaemia experienced in 50% and degree 4.5% of patients (Loibl, 2006). Another study examined 285 patients with operable or locally advanced breast cancer and observed a frequency of anaemia in 94.5% of patients, grade 1 (58.7%); grade 2 (34.2%); and grade 3 (1.6%) (Minckwitz, 2005). It was shown as many as 11% - 42% of patients experience grade II-IV neutropenia which occurs in the 1st to 5th cycle. The incidence of leukopenia also occurs in several patients throughout the cycle with varying degrees of severity ranging from 4% - 50%. It is known that the TAC chemotherapy regimen has been classified as a regimen with a high risk of inducing neutropenia and FN (Ashariati, 2022). A study involving 61 patients receiving a TAC chemotherapy regimen along with peg-filgrastim prophylaxis also reported incidents of chemotherapy-induced neutropenia for at least one cycle. Based on the data obtained the incidence of neutropenia in the high TAC regimen, however, the case of FN must be investigated further because FN has involved fever. Neutropenia often occurs at the beginning of the chemotherapy cycle. Based on Lalami (2017), the risk of FN is greater during the first cycle of chemotherapy because patients usually receive full-dose intensity chemotherapy. The incidence of thrombocytopenia was also found in this study. It was shown as many as 11% - 25% of patients experience grade I thrombocytopenia during the 1st to 3rd cycles.

As many as 25% - 75% of patients experience anaemia that occurs in all cycles of chemotherapy with degrees I-II severity. This is in accordance with a study conducted by Zhu (2015) that the incidence of anaemia of grade 2 or more varies from 18.2% to 59.7% in breast cancer patients treated with docetaxel and cyclophosphamide regimens. Leukopenia and neutropenia also occur in the TC regimen. Grade I neutropenia occurs in 20% - 25% of patients in cycles 1, 4, and 5 and grade IV neutropenia occurs in 20% - 25% of patients in cycles 2, 4, and 3 5. FN is the most common effect of chemotherapy in patients receiving TC regimens. Leukopenia grades I-IV occurs in 20% - 60% of patients in the 1st to 6th cycles. In this study, no thrombocytopenia was found in patients receiving the TC regimen.

The anaemia found in this study occurred in all chemotherapy cycles with grade I-III severity in 8% - 70% of patients. Anaemia in the TCH regimen occurs in 96% of patients with degrees I-IV severity and most patients experience grade II (Zhu, 2015). In this study, the incidence of neutropenia and leukopenia occurred frequently in the TCH regimen in all cycles with grades I-IV. Grade II-IV neutropenia was experienced by 8% - 23% of patients in the first cycle of chemotherapy. Grade I-IV leukopenia was experienced by 8% - 23% of patients in the first cycle of chemotherapy. In another study, patients may experience varying degrees of myelosuppression, and the ratio of grades III-IV neutropenia on the TCH regimen was 25.6% of 39 cases experiencing neutropenia (Chen, 2015). Adverse reactions caused by administration of chemotherapeutic agents include myelosuppression to varying degrees.

## Conclusion

- Based on the study on myelosuppression side effect in breast cancer patients conducted from March to June 2023 at Division of Hematology and Medical Oncology, Airlangga University Hospital, it can be concluded that all breast cancer chemotherapy regimen can cause myelosuppression. The chemotherapy regimens used in breast cancer patients at Airlangga University Hospital are AC-T, TAC, Paclitaxel-Carboplatin, TC, AC-TH, and TCH.
- Anaemia grade I-II occurred in 50% - 100% patients who had AC-T regimen. Neutropenia grade I-IV occurred in 8% - 54% patients who had TCH regimen. Leukopenia grade I-IV occurred in 20% - 60% patients who had TC regimen. Thrombocytopenia grade I occurred in 20% - 33% who Paclitaxel-Carboplatin regimen. The myelosuppression side effect occurred in cycles 1 to 6.
- The incidence of delayed chemotherapy in patients due to myelosuppression is 18%.

## Data Availability

All relevant data are within the manuscript and its Supporting Information files.

## Acknowledgement

The writers of this study like to thank you for all the Division of Hematology-Oncology Airlangga University Hospital staffs for providing data for this study

## Grants and/or funding information

There is no funding for this study.

## Conflict of interest statement

The writers declare there is no conflict of interest for this study.

## Notes

### Competing Interest Statement

The authors have declared no competing interest.

### Funding Statement

The author(s) received no specific funding for this work.

### Author Declarations

The study was approved by the Research Ethics Committee of Rumah Sakit Universitas Airlangga and was declared to have passed the ethical review with a Certificate of the Ethics Review Approval Number 021/KEP/2023

